# Phase 1 Safety and Pharmacokinetics Studies of BRII-196 and BRII-198, SARS-CoV-2 Spike-Targeting Monoclonal Antibodies

**DOI:** 10.1101/2021.07.21.21260964

**Authors:** Yao Zhang, Xiaohua Hao, Ji Ma, Mingming Wang, Yanyan Li, Yang Liu, Dong Zhao, Wen Zhang, Chunming Li, Li Yan, Qing Zhu, Fujie Zhang

## Abstract

**Background:** BRII-196 and BRII-198 are two anti-SARS-CoV-2 monoclonal neutralizing antibodies with modified Fc region that extends half-life and are being developed as cocktail therapy for the treatment of COVID-19. Safety, tolerability, pharmacokinetics, and immunogenicity of BRII-196 and BRII-198 were investigated in healthy adults.

**Methods:** Single ascending doses of BRII-196 and BRII-198 were evaluated in parallel in the first-in-human, placebo-controlled phase 1 studies. A total of 32 healthy adults were randomized and received a single intravenous infusion of 750, 1500, and 3000 mg of BRII-196 (n=12), BRII-198 (n=12), or placebo (n=8) and were followed for 180 days.

**Results:** All infusions were well tolerated at infusion rates between 0.5 mL/min to 4 mL/min with no dose-limiting adverse events, deaths, serious adverse events, or any systemic or local infusion reactions. Most treatment-emergent adverse events were isolated asymptomatic laboratory abnormalities of Grade 1-2 in severity. Each mAb displayed pharmacokinetics expected of Fc-engineered human IgG1 with mean terminal half-lives of approximately 46 days and 76 days, respectively, with no evidence of significant anti-drug antibody development.

**Conclusions:** BRII-196 and BRII-198 were well-tolerated. Clinical results support further development as therapeutic or prophylactic options for SARS-CoV-2 infection.

## Background

As of March 2021, globally, over 120 million people have been infected by SARS-CoV-2, resulting in more than 2.6 million death by Coronavirus Disease 2019 (COVID-19). The number of cases continues to rise[1].

The development of safe and effective therapeutics represents a key strategy to curb COVID-19 mortality and morbidity. Several approved medications (e.g., chloroquine, hydroxychloroquine, lopinavir/ritonavir) have been “repurposed” and evaluated, none of which have been shown associated with improved clinical outcomes[2, 3]. There have been limited treatment options, including remdesivir, dexamethasone, and convalescent plasma, that demonstrated clinical efficacy in subsets of patients with COVID-19[4-7].

Neutralizing monoclonal antibodies (mAbs) represent a promising therapeutic option in preventing and treating known and emerging infectious diseases, including viruses[8]. Recent clinical trials of mAbs reported positive results indicating virological and clinical benefits among outpatients with COVID-19 [9, 10]. In addition, several COVID-19 vaccines offering>90% protection have become available for emergency use [11, 12]. However, recent data showed that several mutations identified in the receptor binding domain (RBD) of spike protein in the emerging new variants conferred reduced activity to neutralization by the antibodies that were developed as a therapy for treatment or elicited by vaccines for prevention [13, 14]. These emergent variants present new challenges for mAb therapy and threaten the protective efficacy of current vaccines and mAbs. Therefore, additional mAb treatment options, especially a cocktail of antibodies targeting distinct epitopes, are needed as a critical strategy to reduce the generation and selection of resistant viruses during treatment and ensures binding and neutralization of the antibodies to new variants.

BRII-196 and BRII-198, two recombinant human IgG1 mAbs that target the distinct epitope regions in RBD in coronavirus spike glycoproteins non-competitively, are under clinical development as a cocktail for the treatment of COVID-19 in outpatients. BRII-196 and BRII-198 are derived directly from human B cells of patients who recovered from COVID-19 [15] with effective neutralizing activity against not only the original isolate of SARS-CoV-2 but also the new variants of concerns, including B.1.351 (as known South African variant) and P1 from Brazil [16, 17]. BRII-196 and BRII-198 are engineered with a triple-amino-acid (M252Y/S254T/T256E [YTE]) substitution in the fragment crystallizable (Fc) region to allow an extended half-life [18, 19] and reduced Fcγ receptor binding for potential risk of Fc-medicated antibody dependent enhancement (ADE) (unpublished data).

The back-to-back first-in-human phase 1 studies in healthy adult participants were designed and conducted to evaluate the safety, tolerability, and pharmacokinetics (PK) of BRII-196 and BRII-198 individually before initiating clinical studies of BRII-196 and BRII-198 in combination in patients.

## Methods

### Study design and participants

BRII-196-001 and BRII-198-001 are two first-in-human phase 1, randomized, single-blind, placebo-controlled, single ascending dose escalation studies in which BRII-196 and BRII-198 were evaluated respectively among healthy adults. These studies were conducted at a single phase 1 unit in China from July 2020 to February 2021 (registered at ClinicalTrials.gov under registration number NCT04479631 and NCT04479644). The study protocol, amendments, and informed-consent forms were reviewed and approved by the Institutional Ethics Committee (IEC). The IEC included Guo’an Wu, Wangyan Jia, Xiulan Li, Xiangmei Zhao, Zhiyun Yang, Xingwang Li, Xianbo Wang, Hanqiu Zhan, Hui Zeng, Yajie Wang, Yun’ao Zhou, and Gang Wan from Beijing Ditan Hospital, Capital Medical University; Kairong Wang and Yangang Tian from Beijing Lvli Law Firm; and Jianguo Sun as a community representative. The study was conducted in accordance with the International Conference on Harmonisation Guidance for Good Clinical Practice guidelines and all applicable local regulatory requirements and laws. Participants provided written informed consent before any study-related procedures were performed.

Inclusion and exclusion criteria for the two studies were identical. Eligible participants were healthy male or female adults aged 18 to 49 years, had a body weight ≤100 kg and a body mass index of 18-24 kg/m^2^, and were in good health determined by no clinically significant findings from medical history, physical examination including vital signs, electrocardiogram (ECG), and clinical laboratory assessments. Women were not pregnant or lactating. Exclusion criteria included a history of SARS-CoV-2 infection or exposure, a history of severe allergic reactions, use of any medications that were started within 14 days before randomization, a positive test for human immunodeficiency virus (HIV) or hepatitis B virus or hepatitis C virus, a history of drug or alcohol abuse within 1 year before the screening, or any history of a medical or psychiatric condition that would place them at risk or interfere with study participation. Participants were also excluded if they were unwilling to use necessary contraception during the study. A complete list of exclusion criteria is included in Supplementary Material 1.

### Study Procedures

Three dose-level cohorts (750, 1500, and 3000 mg) were included to evaluate each mAb and were initiated in a dose-escalation design. In each cohort, participants were randomized at a 3:1 ratio to receive either the single mAb (n=3 or 6) or placebo (n=1 or 2). mAbs or placebo was dispensed into normal saline and was administered via intravenous infusion on Day 1. The infusion started at the initial rate of 0.5 mL/min, gradually increasing to a maximum rate of 4 mL/min. The participants were masked to the treatment assignment and were admitted to the phase 1 unit for monitoring for 7 days post-dosing. The first participant in each cohort who received mAb infusion was monitored for 24 hours before administering study intervention to the subsequent participants. A Safety Review Committee made Dose-escalation recommendations upon review of predefined safety data.

Vital signs, physical examinations, safety laboratory tests including hematology and chemistry, and ECG were obtained before and after study drug administration on days 2, 4, 6, 8, 15, 22, 31, 61, 91, 121, 151, and at the end of the study on day 181. All adverse events (AEs), including infusion reactions and serious adverse events (SAEs) occurring throughout the trial, were recorded and assessed by study physicians at the site. AEs were coded with the Medical Dictionary for Regulatory Activities (MedDRA) version 23.1, and severity was graded using the Common Terminology Criteria for Adverse Events (CTCAE) version 5.0, November 27, 2017.

### Pharmacokinetics and Immunogenicity Assessments

The pharmacokinetics parameters were evaluated using blood samples collected before dosing, at 3 and 8 hours post dose, and days 2, 4, 6, 8, 15, 22, 31, 61, 91, 121, 151, and 181. BRII-196 or BRII-198 serum concentrations were measured using validated enzyme linked immunosorbent assays (ELISA) with the lower limit of quantitation (LLOQ) at 150 ng/mL. Serum samples for anti-drug antibody (ADA) assays were collected at the following time points: pre-dose on day 1 and at days 15, 31, and 181. Two validated titer-based 3-stage ELISA assays were used to detect presences of ADAs from subjects over the study durations.

### Study Outcomes

Primary endpoints were incidence of adverse events (AEs) and change from pre-dose baseline in clinical assessments, including vital signs, ECG readings, and laboratory results. Secondary endpoints included the pharmacokinetics profiles and presence of ADAs to BRII-196/BRII-198 in samples collected after dosing for up to 180 days.

### Statistical Analysis

The sample size of each study was consistent with a phase 1 first-in-human study. The safety analysis included all participants who were randomized and received any dose of the study drug. Categorical and continuous data were summarized descriptively. Participants were analyzed according to the study drug they received. Participants receiving placebo in different cohorts of each study were pooled in the analysis.

The PK parameters were estimated by non-compartmental analyses using WinNonlin module in the Phoenix Platform (version 8.3.1.5014, Certara Inc., Princeton, NJ 08540). Calculations were performed prior to rounding, and nominal sampling times were used in the pharmacokinetic analysis. All pharmacokinetic parameters and summary statistics are reported to 3 significant digits except for T_max_ (reported in median values), which is reported to 1 decimal place.

## Data Availability

The datasets generated during and/or analysed during the current study are available from the corresponding author on reasonable request.

## Results

From July 12, 2020, to August 10, 2020, a total of 135 participants were screened, with 16 and 17 eligible and enrolled for BRII-196-001 and BRII-198-001 studies, respectively (**Figure 1**). The demographics of the study population are summarized in **Table 1**, consisting of 28 (84.8%) men and 5 women with a median age of 33.0 years and a median BMI of 21.9 kg/m^2^.

**Table 1.** Participant Demographics and Baseline Characteristics.

|  | **BRII-196-001** |  |  |  | **BRII-198-001** |  |  |  |
| --- | --- | --- | --- | --- | --- | --- | --- | --- |
|  | **750 mg**<br>**(n=3)** | **1500 mg**<br>**(n=6)** | **3000 mg**<br>**(n=3)** | **Placebo**<br>**(n=4)** | **750 mg**<br>**(n=3)** | **1500 mg**<br>**(n=7)** | **3000 mg**<br>**(n=3)** | **Placebo**<br>**(n=4)** |
| Median age, years | 22.0 | 32.5 | 34.0 | 33.5 | 30.0 | 33.0 | 30.0 | 37.5 |
| Men | 3 (100%) | 4 (67%) | 3 (100%) | 3 (75%) | 2 (67%) | 7 (100%) | 3 (100%) | 3 (75%) |
| Women | 0 | 2 (33%) | 0 | 1 (25%) | 1 (33%) | 0 | 0 | 1 (25%) |
| Race |  |  |  |  |  |  |  |  |
| Han, Chinese | 3 (100%) | 6 (100%) | 2 (67%) | 3 (75%) | 3 (100%) | 6 (86%) | 3 (100%) | 4 (100%) |
| Other, Chinese | 0 | 0 | 1 (33%) | 1 (25%) | 0 | 1 (14%) | 0 | 0 |
| Median body-mass index,<br>kg/m<sup>2</sup> | 22.80 | 21.15 | 21.60 | 22.90 | 23.70 | 21.00 | 22.30 | 23.10 |

**Figure 1.**
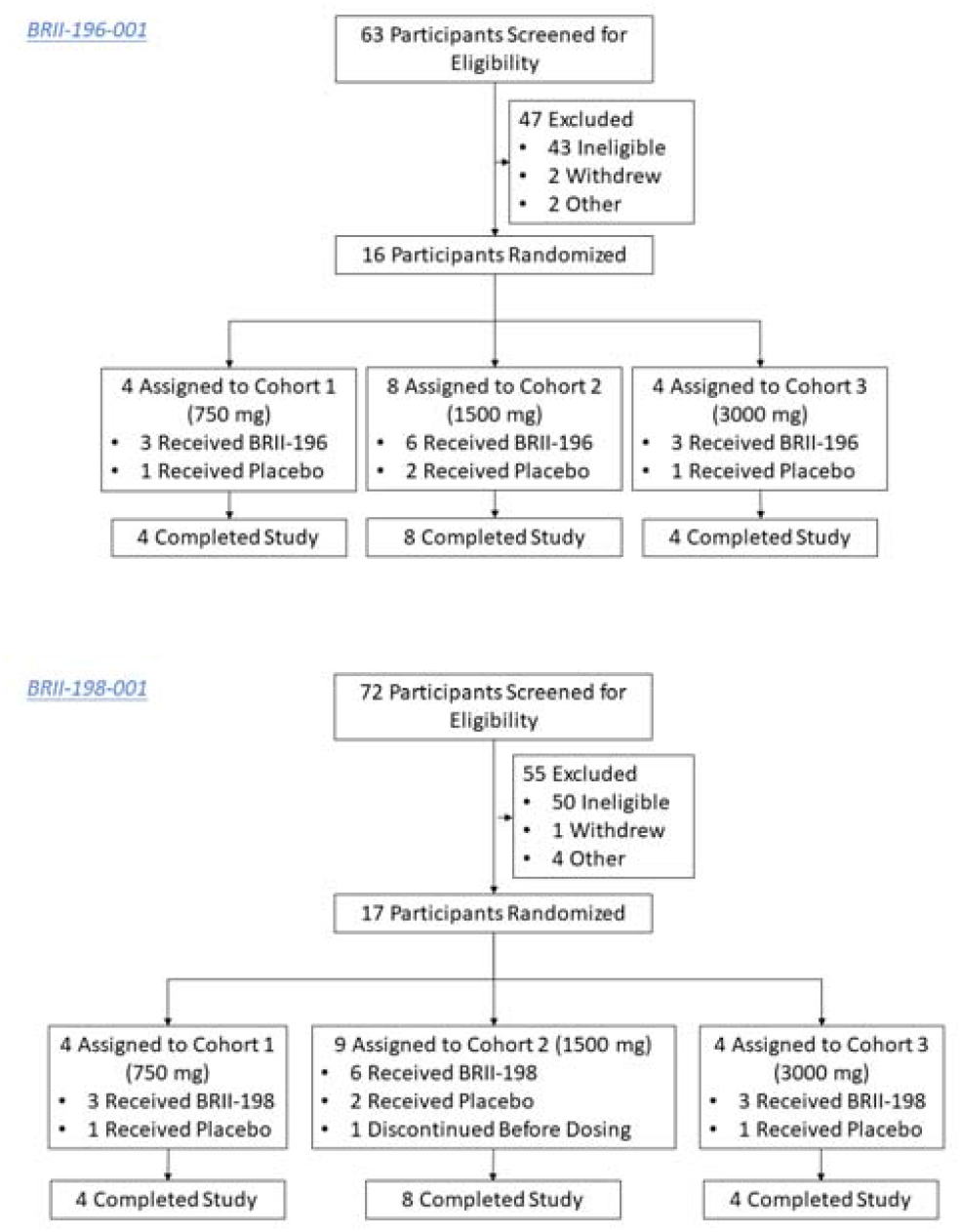
Trial Profiles.

A total of 12 participants received a single dose of BRII-196 IV infusion, 12 participants received a single dose of BRII-198 IV infusion, and 8 participants received placebo (normal saline). One participant randomized in BRII-198-001 study did not receive the infusion because of elevated blood pressure before dosing. All participants received their full planned dose except for 3 participants assigned to 750 mg of BRII-196 and 1 participant assigned to placebo for whom the received volume was approximately 10% less than the intended volume due to the remaining volume left within the infusion set. All participants dosed have completed study visits. Total infusion duration for BRII-196 or BRII-198 at investigated dose levels were 1.53-1.70 hours (750 mg), 2.27-2.43 hours (1500 mg), and 3.58-3.75 hours (3000 mg) (Supplementary Table 1).

### Safety

BRII-196 and BRII-198 administration was safe and well-tolerated. No deaths, serious AEs, treatment-emergent adverse events (TEAEs) leading to early termination, or infusion-related reactions occurred during the study. Adverse events reported in BRII-196-001 and BRII-198-001 are summarized in **Table 2** and **Table 3**, respectively. 26 (81%) participants had at least one treatment-emergent adverse events, including 18 (75%) participants receiving BRII-196 or BRII-198 compared to 8 (100%) participants receiving placebo. Overall incidences of adverse events were balanced between groups receiving BRII-196 or BRII-198 and placebo. The most common TEAEs were isolated asymptomatic laboratory abnormalities of CTCAE Grade 1 or 2 in severity that did not require medical intervention and typically normalized or returned to baseline level within four weeks (Supplementary Table 2). Seven out of 24 participants had 8 Grade 1 or 2 adverse events that were considered by the investigators as related to BRII-196 or BRII-198, including increased blood total bilirubin, increased alanine aminotransferase, decreased neutrophil count, and decreased white blood cell count (Supplementary Table 3). No safety issues were seen with respect to vital signs or ECG measurements. No dose-dependent pattern of any adverse events was observed.

**Table 2.** Summary of Adverse Events of BRII-196-001 Study.

|  | BRII-196<br>750mg<br>(n=3) | BRII-196<br>1500mg<br>(n=6) | BRII-196<br>3000mg<br>(n=3) | Placebo<br>(n=4) |
| --- | --- | --- | --- | --- |
| Participants with any TEAE | 3 (100%) | 5 (83%) | 2 (67%) | 4 (100%) |
| Participants with Grade 1 TEAE | 3 (100%) | 5 (83%) | 2 (67%) | 4 (100%) |
| Participants with Grade 2 TEAE | 1 (33%) | 3 (50%) | 0 | 2 (50%) |
| Participants with Grade 3 or above TEAE | 0 | 0 | 0 | 0 |
| Participants with serious TEAE | 0 | 0 | 0 | 0 |
| Participants with infusion reaction TEAE | 0 | 0 | 0 | 0 |
| Participants with TEAEs related to study drug | 1 (33%) | 3 (50%) | 0 | 0 |
| Participants with any TEAE leading to discontinuation of study drug | 0 | 0 | 0 | 0 |
| Death | 0 | 0 | 0 | 0 |
| Number of participants with AEs by MedDRA SOC <sup>a</sup> |  |  |  |  |
| Investigations | 3 (100%) | 4 (67%) | 2 (67%) | 4 (100%) |
| Infections and infestations | 1 (33%) | 0 | 0 | 2 (50%) |

TEAE=treatment-emergent adverse event. SOC=system organ class.
Participants who experienced the same AE on more than one occasion (based on the specific category) are counted once in each relevant category. Percentages are based on the number of participants in the treatment group.
<sup>a</sup> Only SOC's experienced by $\geq 2$ participant or with the maximum severity category ≥ Grade 2 are included.

**Table 3.** Summary of Adverse Events of BRII-198-001 Study.

|  | BRII-198<br>750mg<br>(n=3) | BRII-198<br>1500mg<br>(n=6) | BRII-198<br>3000mg<br>(n=3) | Placebo<br>(n=4) |
| --- | --- | --- | --- | --- |
| Participants with any TEAE | 3 (100%) | 5 (83%) | 0 | 4 (100%) |
| Participants with Grade 1 TEAE | 2 (67%) | 5 (83%) | 0 | 3 (75%) |
| Participants with Grade 2 TEAE | 1 (33%) | 0 | 0 | 1 (25%) |
| Participants with Grade 3 or above TEAE | 0 | 2<sup>a</sup> (33%) | 0 | 1<sup>b</sup> (25%) |
| Participants with serious TEAE | 0 | 0 | 0 | 1<sup>b</sup> (25%) |
| Participants with infusion reaction TEAE | 0 | 0 | 0 | 0 |
| Participants with TEAEs related to study drug | 2 (67%) | 1 (17%) | 0 | 0 |
| Participants with any TEAE leading to discontinuation of study drug | 0 | 0 | 0 | 0 |
| Death | 0 | 0 | 0 | 0 |
| Number of participants with AEs by SOC <sup>c</sup> |  |  |  |  |
| Investigations | 3 (100%) | 4 (67%) | 0 | 3 (75%) |
| Gastrointestinal disorders | 1 (33%) | 1 (17%) | 0 | 0 |
| Injury, poisoning and procedural complications | 0 | 0 | 0 | 1 (25%) |

TEAE=treatment-emergent adverse event. SOC=system organ class.
Participants who experienced the same AE on more than one occasion (based on the specific category) are counted once in each relevant category. Percentages are based on the number of participants in the treatment group.
<sup>a</sup> 1 participant had an elevated blood creatine phosphokinase of Grade 4 after excessive exercise, and 1 participant had an increased blood triglycerides of Grade 3 on Day 181. Both events were not considered as related to the study drug by the investigators and normalized within 1-3 weeks.
<sup>b</sup> 1 participant receiving placebo had a serious TEAE (bilateral traumatic foot fracture leading to hospitalization, which was also reported as a Grade 3 TEAE). The participant recovered after surgery.
<sup>c</sup> Only SOCs experienced by $\geq 2$ participant or with the maximum severity category ≥ Grade 2 are included.

### Pharmacokinetics

Pharmacokinetic data were available for a total of 12 participants each for BRII-196 and BRII-198 over the 181-day study duration. Following a single intravenous infusion of BRII-196 or BRII-198 at 750, 1500, and 3000 mg, mean serum PK parameters, including C_max_ and AUC, increased in an approximately dose-proportional manner (**Table 4, Figure 2**). The mean systemic serum clearances were 75.4 and 57.0 mL/day for BRII-196 and BRII-198, respectively. The mean terminal half-lives (t_1/2_) of BRII-196 and BRII-198 were 44.6-48.6 days and 72.2-83.0 days. The relatively shorter terminal half-life of BRII-196 correlated with slightly higher systemic clearance.

**Table 4.**
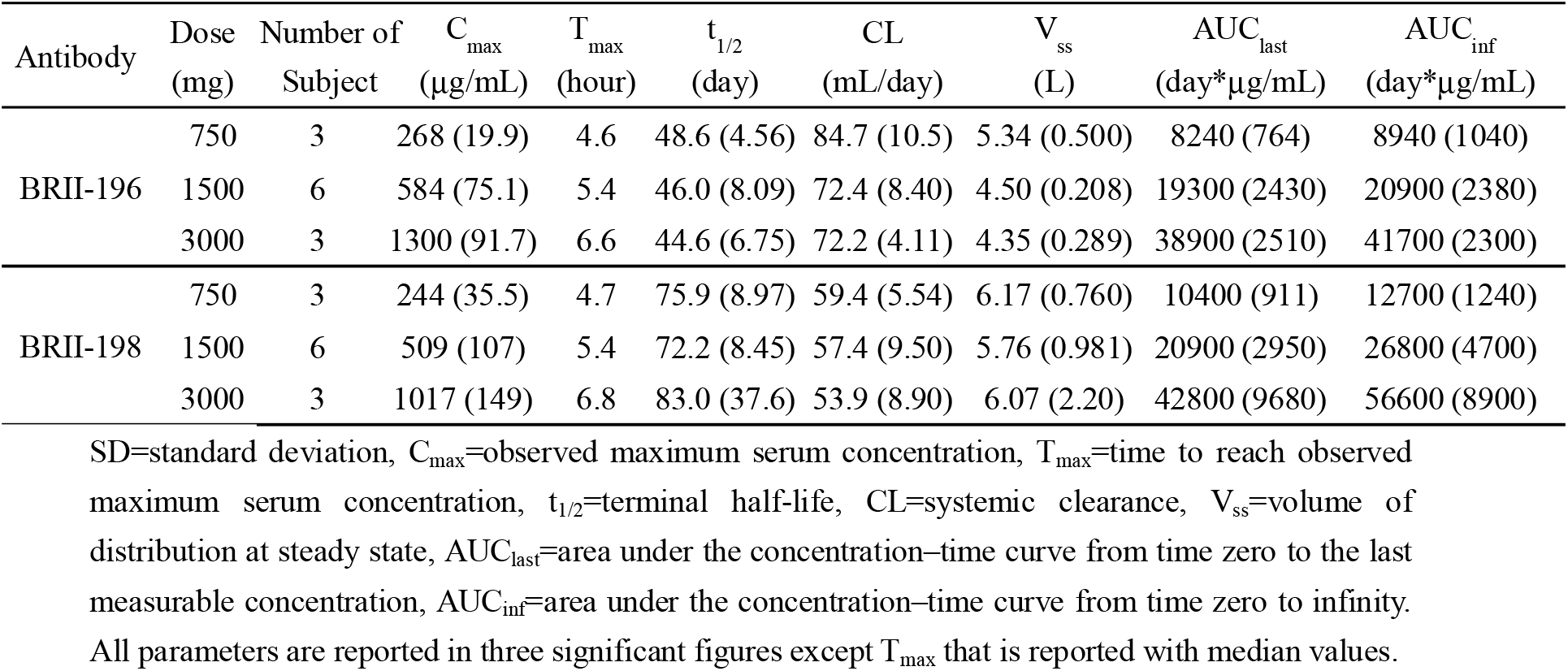
BRII-196 and BRII-198 Pharmacokinetic Parameters Following A Single Intravenous Infusion Administration to The Healthy Adult Participants.

**Figure 2.**
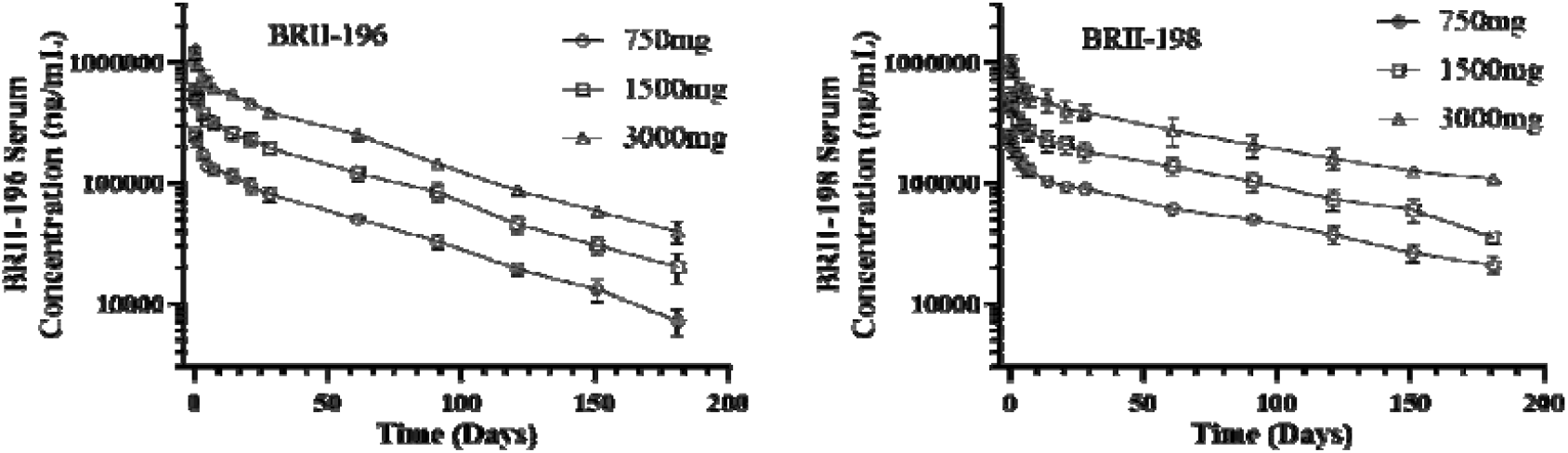
Observed Mean (±Standard Deviation) Serum Concentration - Time Profiles of BRII-196 and BRII-198 Following A Single Intravenous Infusion Administration to The Healthy Adult Subjects.

### ADA Response

In BRII-196-001 study, a total of 4 out of 12 participants had positive ADA samples in the screening assay, but all tested negative in the follow-up confirmatory assay. In BRII-198-001 study, one participant who received BRII-198 had positive ADA samples in the screening assay but later confirmed negative in the tier-2 confirmatory assay.

## Discussion

BRII-196 and BRII-198 are fully human mAbs that bind to noncompeting epitope regions on the RBD of SARS-CoV-2 S glycoprotein that interacts with the angiotensin-converting enzyme 2 (ACE2) to enter human cells [20]. Therefore, they neutralize virus infection by preventing virus binding to ACE2, blocking virus entry and subsequent virus infection as demonstrated in vitro and in vivo [15, 21]. The combined use of 2 independently neutralizing antibodies is likely to reduce the risk of treatment failure from the development of virus escape mutants or new variants that may arise during an outbreak. Currently approved COVID -19 vaccines were constructed against the original virus, of which attenuation of serum neutralization activity has been observed[14, 22]. In a recent trial conducted in South Africa, the ChAdOx1 nCoV-19 vaccine did not protect against mild-to-moderate COVID-19 due to the B.1.351 variant [23]. Pseudo viruses harboring mutations found in B.1.351 South Africa origin, P.1 Brazil origin, B.1.526 New York origin, L452R, and the spike protein from the California origin variant lineage B.1.427/B.1.429, had significantly reduced susceptibility to bamlanivimab [24], a mAb authorized for emergency use. In addition, its approved cocktail with etesevimab under EUA was also tested inactive against the live virus of B.1.351 South Africa [17]. As some of these variants are beginning to spread around the world, it has posed a real threat to vaccine and antibody therapies. By contrast, BRII-196 and BRII-198 cocktail retained their neutralizing activity against all variants tested, including B1.1.7 UK variant, B1.357 South Africa variant, and P1 Brazilian variants[16, 17], thereby could provide more treatment options to patients.

In these first-in-human clinical studies, intravenous administration of BRII-196 and BRII-198 were both safe and well-tolerated in healthy adult participants receiving a single dose up to 3000 mg, with no infusion reactions or AEs leading to adjustment or discontinuation of the infusion. Study participants were followed up to 180 days post-dosing considering the expected prolonged half-lives of the mAbs. Except for one SAE, one Grade 4 elevation of creatine phosphokinase, and one Grade 3 elevation of triglycerides, all deemed as not related to study drugs, all AEs reported were Grade 1-2 in severity, with the majority being isolated asymptomatic transient laboratory abnormalities. There were no safety concerns regarding vital signs or ECGs. The safety observations in these phase 1 studies are consistent with the expected safety profile of human IgG1 mAbs in healthy subjects with no endogenous target.

With YTE modification, both BRII-196 and BRII-198 demonstrated anticipated 2-3 folds half-life extension in healthy adult subjects. In comparison with BRII-198, the relatively shorter half-life observed with BRII-196 correlates with slightly higher systemic clearance. As both antibodies showed comparable increased binding affinities to human and cynomolgus monkey FcRns at pH 6.0 in vitro (unpublished data), different molecular properties such as charge variants, glycan profiles, and overall thermal stability might contribute to the observed differences. With up to a 3-week optimal treatment window for neutralizing antibodies for COVID-19 patients, both antibodies are expected to provide comparable target coverage at the same dose level.

BRII-196 and BRII-198 have several advantages that may facilitate their use for the treatment and prevention of SARS-COV-2 infections. First, mAbs targeting specific viral epitopes, including BRII-196 and BRII-198, belong to a platform technology associated with a favorable safety profile. Findings regarding safety and tolerability in phase 1 trials of BRII-196 and BRII-198 are consistent with the expectation, which support an expedited development towards approval for future clinical use. Second, targeting distinct viral epitopes in the RBD of SARS-COV-2 spike protein non-competitively, BRII-196 and BRII-198 can be used in combination to provide additive antiviral effect, reduce the generation of resistant viruses to treatment, and retain activity against emergent naturally occurring variants as mentioned above. Third, BRII-196 and BRII-198 are engineered with YTE substitution in the Fc region to allow an extended half-life, demonstrated in the pharmacokinetic findings from both phase 1 studies, with which BRII-196 and BRII-198 can potentially maintain an adequate drug coverage over an extended period of time and subsequently are better positioned for prophylactic use. Lastly, while YTE improves the half-life of the antibody, it also reduced binding activity of BRII-196 and BRII-198 against human Fcγ receptors, thereby minimizing the potential risk of Fc-mediated antibody dependent enhancement of diseases.

In summary, BRII-196 and BRII-198 were generally safe and well-tolerated in healthy adult participants. Pharmacokinetic and immunogenicity assessments demonstrated linear pharmacokinetics with prolonged half-lives and no ADA development up to 180 days after a single dose. These findings support further development of BRII-196 and BRII-198 for the treatment and prophylaxis of SARS-COV-2 infection in the pandemic outbreak. BRII-196 and BRII-198 combination is currently under development in ACTIV-2, a platform phase 2/3 clinical study, for treatment of COVID-19 in outpatients [25].

## Data Availability

The datasets generated during and/or analysed during the current studies are available from the corresponding author on reasonable request.

## Acknowledgment

The authors thank all the study participants in these clinical studies, without whom the studies would not have been possible. We thank Dr Yibo Zhou, Jianmin Jing, and Yonghong Mang, for their contributions in the studies. We also thank D.Margolis for criticically reviewing the manuscript and Dr. Chan Gao for her assistance with formatting/editing and submission.

## Financial support

These trials were sponsored by TSB Therapeutics, a subsidiary company of Brii Biosciences. Y.Z., J.M., Y.L., Y.Y.L., M.W., C.L., L.Y., and Q.Z. are employees of Brii Biosciences. Y.Z. and Q.Z. are employees of TSB Therapeutics. F.J.Z. received support from TSB for participation as a principal investigator in the study.

## Author contributions

Brii Biosciences was involved in the study design, collection, analysis and interpretation of data, the writing of this report, and the decision to submit this manuscript for publication. F.J.Z., X.H.H, D.Z., and W.Z. reviewed protocol, and were responsible for study implementation and data collection at the study site. All authors at Brii Bioscienecs had access to all study data for review, participated in interpretation of the data and in drafting and revising the article for important intellectual content, and provided final approval of the submitted manuscript. All authors assume responsibility for the integrity and completeness of the reported data. Y.Z., J.M., Y.L., and Q.Z prepared the manuscript.

## Potential conflicts of interest

Y.Z., J.M., Y.L., Y.L., M.W., C.L., L.Y., and Q.Z. are employees of Brii Biosciences. Y.Z. and Q.Z. are employees of TSB Therapeutics. All other authors report no potential conflicts. All authors have submitted the ICMJE Form for Disclosure of Potential Conflicts of Interest.

## Notes

### Clinical Trial

NCT04479631/NCT04479644

### Author Declarations

The study protocol, amendments, and informed-consent forms were reviewed and approved by the Institutional Ethics Committee (IEC). The IEC included Guoan Wu, Wangyan Jia, Xiulan Li, Xiangmei Zhao, Zhiyun Yang, Xingwang Li, Xianbo Wang, Hanqiu Zhan, Hui Zeng, Yajie Wang, Yunao Zhou, and Gang Wan from Beijing Ditan Hospital, Capital Medical University; Kairong Wang and Yangang Tian from Beijing Lvli Law Firm; and Jianguo Sun as a community representative. The study was conducted in accordance with the International Conference on Harmonisation Guidance for Good Clinical Practice guidelines and all applicable local regulatory requirements and laws. Participants provided written informed consent before any study-related procedures were performed.

