## Supplemental Material for "Phase 1 Safety and Pharmacokinetics Studies of BRII-196 and BRII-198, SARS-CoV-2 Spike-Targeting Monoclonal Antibodies"

#### Supplementary material

#### Inclusion and Exclusion Criteria for BRII-196-001 and BRII-198-001 Studies

### Inclusion Criteria

Subjects are eligible to be included in the study only if all of the following criteria apply:

Age

1. Subject must be 18 to 49 years of age inclusive, at the time of signing the informed consent.

Type of Subjects

2. Subjects who are healthy as determined by medical evaluation including medical history, full physical examination, laboratory tests, and cardiac monitoring using 12-lead ECG, and have venous access sufficient to allow for blood sampling as per the protocol.

Weight

3. Body weight ≤100 kg and body mass index (BMI) within the range of 19.0-24.0kg/m^2^ (inclusive). The weight of male subjects shall be no less than 50kg, and that of female subjects shall be no less than 45kg.

Sex

4. Male and female

Contraceptive use by men or women should be consistent with local regulations regarding the methods of contraception for those participating in clinical studies.

a. Male subjects:
   i. Male subjects with female partners of childbearing potential must agree to meet one of the following contraception requirements from the time of study agent administration through the end of the study:

- total abstinence, or
- vasectomy with documentation of azoospermia, or
- barrier form of contraception with spermicide, or
- female partner use of an intrauterine device or hormonal contraceptives including oral, implantable, injectable or transdermal contraceptives within 30 days prior to male subject signing the informed consent form (ICF).
  ii. Male subjects must also agree not to donate sperm through the end of the study.

b. Female subjects:
i. A female subject is eligible to participate if she is:

- of non-childbearing potential, defined as premenarchal, or pre-menopausal with a documented hysterectomy, or bilateral tubal ligation or oophorectomy.
- of childbearing potential, not breastfeeding and agrees to use one of the following acceptable methods of contraception through the end of the study:
- total abstinence, or
- barrier form of contraception such as condom or occlusive cap with spermicide, or
- an intrauterine device, or hormonal contraceptives including oral, implantable, injectable or transdermal contraceptives initiated at least within 30 days prior to signing the ICF.

ii. Woman of Childbearing Potential* **(**WOCBP) must have a negative serum pregnancy test at screening and a negative serum pregnancy test on Day 1 prior to study treatment (within 24 hours) administration.

**Informed** **Consent**

5. Capable of giving signed informed consent as described in the protocol which includes compliance with the requirements and restrictions listed in the ICF and in this protocol.

**Other Inclusions**

6. Agrees to not donate blood during the duration of the study.
7. Agrees not to increase physical activity for 4 weeks after study agent administration.

### Exclusion Criteria

Subjects are excluded from the study if any of the following criteria apply:

**Medical Conditions**

1. Significant history or clinical manifestation of any metabolic, allergic, dermatological, hepatic, renal, hematological, pulmonary, cardiovascular, gastrointestinal, neoplastic (with the exception of basal or squamous cell cancer), neurological, or psychiatric disorder (as determined by the Investigator) capable of significantly altering the absorption of drugs; of constituting a risk when taking the study intervention; or of interfering with the interpretation of the data.
2. SARS-COV-2 exposure history or clinical history of COVID-19, including positive COVID-19 RNA test one week before screening or at screening.
3. History of previous diagnosis of SARS.
4. Any history of a severe allergic reaction with generalized urticaria, angioedema, or anaphylaxis prior to enrollment that has a reasonable risk of recurrence during the study.
5. A history of significant hypersensitivity, intolerance, or allergy to any drug compound, food, or other substance, unless approved by the Investigator.
6. Bleeding disorder diagnosed by a doctor (e.g., factor deficiency, coagulopathy, or platelet disorder requiring special precautions) or significant bruising or bleeding difficulties with blood draws.
7. Any major surgical procedure or hospitalization within 3 months prior to Day 1 or during the study, unless deemed not clinically significant by the Investigator.
8. History or presence of an abnormal ECG which, in the Investigator's opinion, is clinically significant.
9. History of alcohol or other substance abuse within 1 year prior to screening according to the Diagnostic and Statistical Manual of Mental Disorders, 5th edition criteria, or recent use of drugs of abuse or a positive urine screen for drugs of abuse at screening.
10. History of depression or suicidal thoughts and/or behaviors within 1 year prior to screening.
11. History of clinically significant chronic liver disease from any cause, presence of hepatitis B surface antigen, hepatitis C virus antibody, human immunodeficiency virus antibody, or syphilis antibody.

**Prior/Concomitant Therapy**

12. Use prescription drugs within 14 days before Day 1, or expected in the course of the study.
13. Use of antiplatelet, anticoagulant, or antiepileptic medications within 30 days prior to Day 1, or expected in the course of the study.
14. Received immunoglobulin or blood products in the 6 months prior to Day 1, or expected in the course of the study.
15. Received any vaccine within 14 days prior to Day 1 through 180 days after dosing in the study.
16. Use of over the counter (OTC) medication or herbal remedy, e.g., Traditional Chinese Medicine, within 14 days before Day 1, or expected in the course of the study.

**Prior/Concurrent Clinical Study Experience**

17. Received an investigational agent or provided clinical samples for clinical studies within 90 days or 5 half-lives of the investigational agent whichever is longer, before screening or are active in the follow-up phase of another clinical study involving interventional treatment. Subjects must also agree not to take part in any other study at any time during their participation in this study, inclusive of the follow-up period.
18. Previous receipt of a licensed or investigational monoclonal antibody within 5 half-lives or 6 months prior to screening, whichever is longer.

**Diagnostic assessments**

19. Systolic blood pressure >140 mmHg or a diastolic blood pressure of >90 mmHg after approximately 10 minutes resting at screening.
20. Calculated glomerular filtration rate of <60 mL/min/1.73m^2^ by estimated glomerular filtration rate (eGFR) using standardized local clinical methodology.

**Other Exclusions**

21. Has an average weekly alcohol intake that exceeds 21 units of alcohol per week (males) or 14 units per week (females) within 30 days prior to screening. One unit: 1 glass of wine 5 oz or 150 mL; 12 oz or 360 mL of beer; 1.5 oz or 45 mL of distilled spirits.
22. Are unwilling to stop alcohol consumption within 72 hr prior to Day 1 (as confirmed by alcohol breath screen) or expected for the 24 hr post-dose period.
23. Need special dietary restrictions, unless the restrictions are approved by the Investigator and/or Brii Biosciences.
24. Any conditions which, in the opinion of the Investigator, would make the subject unsuitable for enrollment or could interfere with the subject’s participation in or completion of the study.
25. Donated blood within 90 days before screening.

**Definition of Woman of Childbearing Potential (WOCBP)**

A woman is considered fertile following menarche and until becoming post-menopausal unless permanently sterile (see below).

If fertility is unclear (e.g., amenorrhea in adolescents or athletes) and a menstrual cycle cannot be confirmed before first dose of study intervention, additional evaluation should be considered.

Women in the following categories are not considered WOCBP

1. Premenarchal
2. Premenopausal female with 1 of the following:

- Documented hysterectomy
- Documented bilateral salpingectomy
- Documented bilateral oophorectomy

For individuals with permanent infertility due to an alternate medical cause other than the above, (e.g., mullerian agenesis, androgen insensitivity), Investigator discretion should be applied to determining study entry.

Note: Documentation can come from the site personnel’s: review of the subject’s medical records, medical examination, or medical history interview.

3. Postmenopausal female

- A postmenopausal state is defined as no menses for 12 months without an alternative medical cause.
- A high follicle stimulating hormone (FSH) level in the postmenopausal range may be used to confirm a postmenopausal state in women not using hormonal contraception or hormonal replacement therapy (HRT).
- Females on HRT and whose menopausal status is in doubt will be required to use one of the non-estrogen hormonal highly effective contraception methods if they wish to continue their HRT during the study. Otherwise, they must discontinue HRT to allow confirmation of postmenopausal status before study enrollment.
