## Supplemental Tables 1-3 for "Phase 1 Safety and Pharmacokinetics Studies of BRII-196 and BRII-198, SARS-CoV-2 Spike-Targeting Monoclonal Antibodies"

### Supplementary Tables

**Supplementary Table 1. Infusion Related Parameters**

| Antibody | Dose | Number of Participants | Total Volume Infused  (ml, median [min, max]) | Total Infusion Duration  (h, median [min, max]) |
| --- | --- | --- | --- | --- |
| BRII-196 | 750 mg | 3 | 114 (110, 116) | 1.57 (1.53, 1.57) |
|  | 1500 mg | 6 | 250 (250, 250) | 2.32 (2.27, 2.43) |
|  | 3000 mg | 3 | 500 (500, 500) | 3.60 (3.58, 3.70) |
|  | Placebo | 4 | 250 (125, 500) | 2.30 (1.55, 3.65) |
| BRII-198 | 750 mg | 3 | 125 (125, 125) | 1.67 (1.65, 1.70) |
|  | 1500 mg | 6 | 250 (250, 250) | 2.33 (2.28, 2.43) |
|  | 3000 mg | 3 | 500 (500, 500) | 3.73 (3.70, 3.75) |
|  | Placebo | 4 | 250 (125, 500) | 2.32 (1.67, 3.73) |

**Supplementary Table 2. Adverse Events by System Organ Class and Preferred Term**

| **Antibody** | **System Organ Class**  **Preferred Term** | **750 mg (n=3)** | **1500 mg (n=6)** | **3000 mg (n=3)** | **Placebo (n=4)** |
| --- | --- | --- | --- | --- | --- |
| **BRII-196** | Investigations | 3 (100%) | 4 (67%) | 2 (67%) | 4 (100%) |
|  | White blood cell count decreased | 1 (33%) | 3 (50%) | 0 | 1 (25%) |
|  | Blood triglycerides increased | 1 (33%) | 0 | 1 (33%) | 2 (50%) |
|  | Neutrophil count decreased | 1 (33%) | 2 (33%) | 0 | 1 (25%) |
|  | Blood uric acid increased | 1 (33%) | 1 (17%) | 1 (33%) | 0 |
|  | Alanine aminotransferase increased | 1 (33%) | 0 | 0 | 1 (25%) |
|  | Blood bilirubin increased | 0 | 1(17%) | 1 (33%) | 0 |
|  | Low density lipoprotein increased | 0 | 0 | 0 | 1 (25%) |
|  | Lymphocyte percentage decreased | 0 | 1 (17%) | 0 | 0 |
|  | Monocyte count decreased | 0 | 0 | 0 | 1 (25%) |
|  | Neutrophil count increased | 0 | 1 (17%) | 0 | 0 |
|  | Platelet count increased | 1 (33%) | 0 | 0 | 0 |
|  | White blood cells urine positive | 0 | 0 | 0 | 1 (25%) |
|  | Infections and infestations | 1 (33%) | 0 | 0 | 2 (50%) |
|  | Nasopharyngitis | 1 (33%) | 0 | 0 | 1 (25%) |
|  | Periodontitis | 1 (33%) | 0 | 0 | 0 |
|  | Tooth infection | 0 | 0 | 0 | 1 (25%) |
|  | Gastrointestinal disorders | 0 | 1 (17%) | 0 | 0 |
|  | Diarrhoea | 0 | 1 (17%) | 0 | 0 |
|  | Musculoskeletal and connective tissue disorders | 0 | 1 (17%) | 0 | 0 |
|  | Pain in extremity | 0 | 1 (17%) | 0 | 0 |
| **BRII-198** | Investigations | 3 (100%) | 4 (67%) | 0 | 3 (75%) |
|  | Blood triglycerides increased | 0 | 4 (67%) | 0 | 2 (50%) |
|  | Blood bilirubin increased | 1 (33%) | 2 (33%) | 0 | 0 |
|  | White blood cell count decreased | 1 (33%) | 1 (17%) | 0 | 1 (25%) |
|  | Alanine aminotransferase increased | 1 (33%) | 1 (17%) | 0 | 0 |
|  | Blood creatine phosphokinase increased | 0 | 1 (17%) | 0 | 0 |
|  | Blood uric acid increased | 0 | 0 | 0 | 1 (25%) |
|  | Aspartate aminotransferase increased | 0 | 1 (17%) | 0 | 0 |
|  | Injury, poisoning and procedural complications | 0 | 0 | 0 | 1 (25%) |
|  | Foot fracture | 0 | 0 | 0 | 1 (25%) |
|  | Ear and labyrinth disorders | 0 | 0 | 0 | 1 (25%) |
|  | Cerumen impaction | 0 | 0 | 0 | 1 (25%) |
|  | Gastrointestinal disorders | 1 (33%) | 1 (17%) | 0 | 0 |
|  | Abdominal pain upper | 1 (33%) | 0 | 0 | 0 |
|  | Constipation | 0 | 1 (17%) | 0 | 0 |
|  | Infections and infestations | 0 | 0 | 0 | 1 (25%) |
|  | Otitis externa | 0 | 0 | 0 | 1 (25%) |
|  | Musculoskeletal and connective tissue disorders | 0 | 0 | 0 | 1 (25%) |
|  | Arthralgia | 0 | 0 | 0 | 1 (25%) |

Participants who experienced the same AE on more than one occasion (based on the specific category) are counted once in each relevant category. Percentages are based on the number of participants in the treatment group.

**Supplementary Table 3. Treatment-related Adverse Events by Preferred Term.**

| **Antibody** | **Preferred Term** | **750 mg (n=3)** | **1500 mg (n=6)** | **3000 mg (n=3)** | **Placebo (n=4)** |
| --- | --- | --- | --- | --- | --- |
| **BRII-196** | White blood cell count decreased | 0 | 1 (17%) | 0 | 0 |
|  | Neutrophil count decreased | 0 | 1 (17%) | 0 | 0 |
|  | Alanine aminotransferase increased | 1 (33%) | 0 | 0 | 0 |
|  | Blood bilirubin increased | 0 | 1 (17%) | 0 | 0 |
| **BRII-198** | Alanine aminotransferase increased | 1 (33%) | 0 | 0 | 0 |
|  | Blood bilirubin increased | 1 (33%) | 1 (17%) | 0 | 0 |

Participants who experienced the same AE on more than one occasion (based on the specific category) are counted once in each relevant category. Percentages are based on the number of participants in the treatment group.
